# Who Is Hospitalized With Whom? Inpatient Contact Networks and Mixing Patterns

**DOI:** 10.1101/2022.02.22.22271374

**Authors:** Kaniz Fatema Madhobi, Ananth Kalyanaraman, Deverick J. Anderson, Elizabeth Dodds-Ashley, Rebekah W. Moehring, Eric T. Lofgren

**Author notes:** Eric T. Lofgren, Paul G. Allen School for Global Health, Washington State University, 240 SE Ott Road, Room 311, Pullman, WA 99164-7090.

## Abstract

**Importance:** Person-to-person contact is important for the transmission of healthcare-associated pathogens. Quantifying these contact patterns is crucial for modeling disease transmission and understanding routes of potential transmission.

**Objective:** Generate and analyze the mixing matrices of hospital patients based on their contacts within hospital units.

**Design, Setting, and Participants:** The study was conducted in 24 hospitals in the Southeastern United States that were part of the Duke Antimicrobial Stewardship Outreach Network (DASON) between January 2015 and December 2017. There were a total of 1,569,413 patients and 299 hospital units.

**Main Outcome and Measures:** The mixing matrices of patients for each hospital unit using age, Elixhauser Score, and a measure of antibiotic exposure.

**Results:** Mixing matrices were calculated from a database of 24 hospitals, which included 2.9 million admission records for nearly 1.6 million patients. Some units had highly similar patterns across multiple hospitals although the number of patients might vary to a great extent. Within a period of 26 months (October 2015 and December 2017), the highest daily average is 765 patients in the ED of Hospital-12 and lowest daily average is only 2 patients in some of the smaller hospital units. For most of the adult inpatient units, frequent mixing was observed for older adult groups while outpatient units e.g. ED and Behavioral Health etc. units showed mixing between different age groups. From the mixing matrices by Elixhauser Score, we observed mixing between patients with relatively higher comorbidity index on the ICUs. Mixing matrices by Antibiotic Rank, a 4-point scale based on priority for antibiotic stewardship programs, resulted in six major distinct patterns due to the variation of the type of antibiotics used in different units.

**Conclusions and Relevance:** The mixing patterns of patients both within and between hospitals followed broadly expected patterns, though with a considerable amount of heterogeneity. These patterns can be used to evaluate the appropriateness of policies and guidelines for smaller community hospitals, as well as improve the design of interventions that rely on altering patient contact patterns.

**Key Points:** *Question:* What are the mixing patterns among hospitalized patients who could be susceptible to infection?

*Finding:* In this study of 299 hospital units from 24 hospitals, we analyzed the mixing patterns between patients based on a number of variables namely Age, Antibiotic Ranks (4-point scale based on priority for antibiotic stewardships programs) and Elixhauser Comorbidity Score. While some units showed highly similar patterns across hospitals, variation has also been observed in concentration of mixing on different age groups and antibiotic usage among the patients who are coming into contact.

*Meaning:* How patients mix, can impact their risk of acquiring an infection. While patterns followed what was expected heuristically, there is considerable between-hospital heterogeneity, which can help in risk assessments and modeling approaches.

## Introduction

An individual’s risk of acquiring an infectious disease is inherently a function of whom they contact, with currently infected individuals representing the exposure source for those individuals infected in the future, a phenomenon known as “dependent happenings”^1^. Therefore, understanding whom an individual contacts becomes critical for understanding their risk. One way of representing and studying this information is the development of a contact network represented by a population of individuals (nodes) and the contacts between them (edges), and studying the properties of this network (i.e. are particular types/classes of people more likely to come into contact with one another than others).

Contact patterns in infectious diseases have been extensively studied in HIV and other sexually transmitted diseases^2–5^, and are being increasingly studied in infectious diseases more broadly^6–9^. Hospitalized patients represent a particularly challenging population for contact network analyses due to the complexity of the hospital environment. Patients may or may not be contacting each other directly (depending on whether they are mobile and can interact with one another), but they may be exposed to pathogens through contamination on the hands and clothes of healthcare workers, on shared instruments, or persisting in the hospital environment as fomites. Several studies have collected hospital contact networks using a variety of methods^10–13^. Many, however, were limited to a single hospital or a single study site. Long-term and multi-site studies of these networks may be important for understanding how hospitals adjust to shifting demands for patient care (e.g. during a pandemic), the evolution of antibiotic stewardship programs, or other shifts to the flow of hospitalized patients, and how this in turn impacts infection control.

The focus of this study was to better describe the contact networks of hospitalized patients using a large, multi-hospital sample. By forming these contact networks, we aimed to visualize contact patterns of variables that affect susceptibility to hospital acquired infections and multidrug resistant organisms (MDRO): age, comorbidity, hospital unit type, and antibiotic exposure. We examine age because it is a known risk factor for infectious diseases such as COVID-19^14^ as well as a number of healthcare-associated infections^15–17^. We also examine Elixhauser score^18^ for comorbidity as a proxy for overall vulnerability to infection, and antibiotic usage as a measure for potential multidrug resistant organism (MDRO) colonization pressure from other patients within the unit^19^.

## Methods

### Patient Data

To estimate the patient contact networks, we used data from the Duke Antimicrobial Stewardship Outreach Network (DASON)^20^ and Duke Health System which contained curated hospital encounter records using uniform definitions for 24 community hospitals and one academic medical center in the Southeastern United States between October 2015 and December 2017. This data is made up of 299 total units across all the hospitals, ranging from 4 units at the smallest hospital to 30 units at the largest. Table 1 shows the basic statistics of the DASON data and corresponding demographics. The Duke University Health System IRB determined this research was exempt from human subjects approval.

**Table 1.**
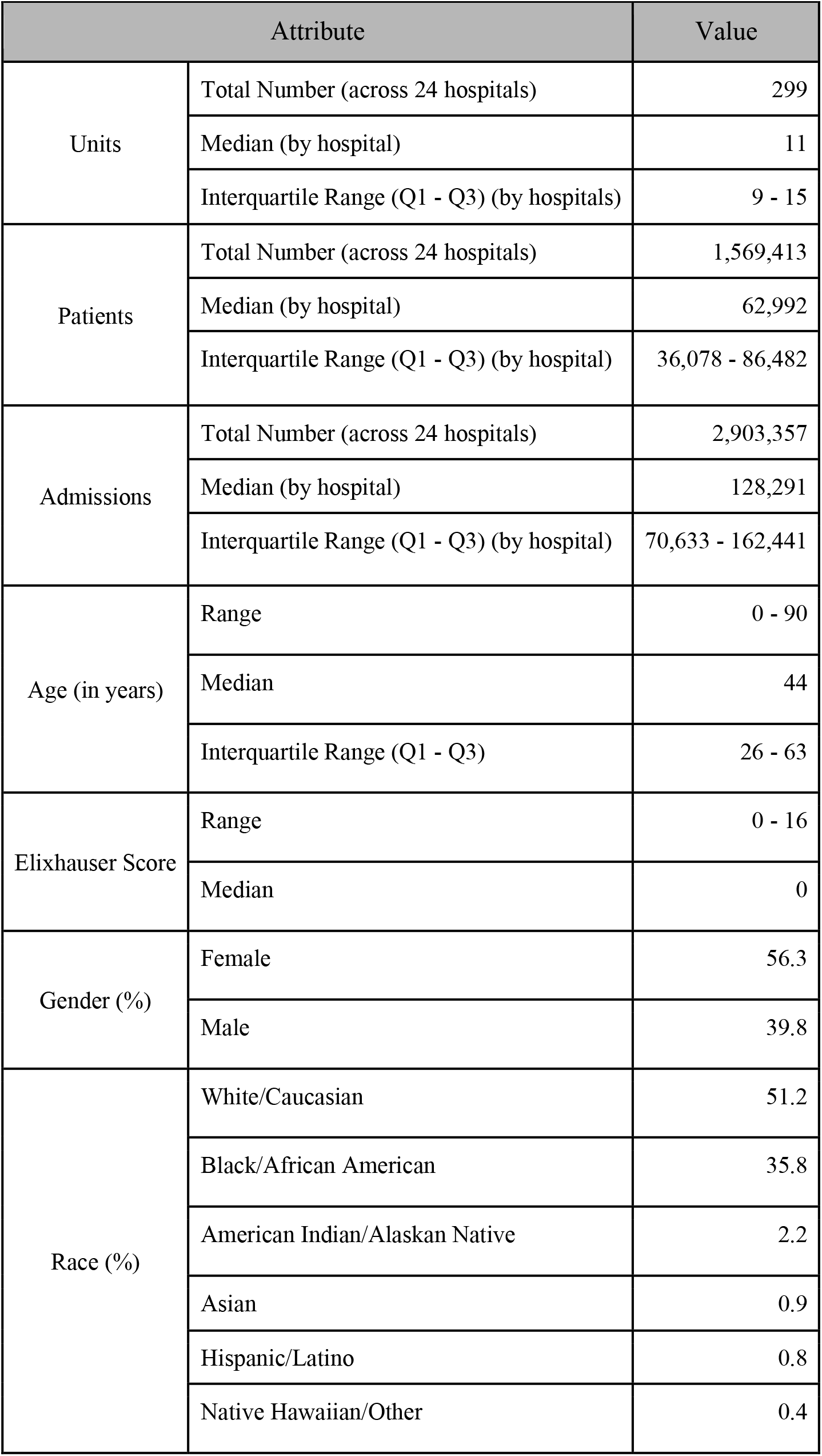
Basic statistics of the DASON database used in our study and corresponding demographic information.

### Network Estimation

Within the DASON data there are records for a patient’s movement between units, as well as arrival and discharge times. Using this information, we estimated a colocation contact network - i.e., if two patients were recorded as being in the same hospital unit during a period of one day, they were counted as being in contact for that day. If there are *k* patients in a unit in a specific day, the number of contacts would be *kC2*. For example, four patients {A, B, C, D} located in the same unit on a specific day will imply six (i.e., *4C2*) pairwise contacts: (A, B), (A, C), (A, D), (B,C), (B, D), (C, D). It is important to note that the *type* of unit is based on its NHSN classification. This classification provides a useful, but ultimately imperfect, approximation of patient case mix, as patients may be placed in a unit for other reasons such as bed availability and hospital volume, a unit’s definition may be shifting over time, etc.

### Computation of Mixing Matrices

Using the pairwise contact information, we computed three types of mixing matrices, based on patient age, Elixhauser score, and antibiotic agent exposure. These mixing matrices record the frequency of contacts between patients belonging to different classes of that category.

For analysis of mixing by age, we construct a two-dimensional table where the patient age(e.g., [0, 90]) is represented by the rows and columns; and cell *(i,j)* corresponds to the number of patient contacts between a patient of age *i* with a patient of age *j*.

Elixhauser Score or Elixhauser Comorbidity Index is a measure of patients comorbidity, developed in 1998^18^. In DASON data, the Elixhauser Score is ranged from [0, 16] where a higher score indicates a greater degree of comorbid conditions in a patient. The mixing matrices in this case are 16×16 tables where the value in a cell [i, j] corresponds to the frequency of contact between a patient having comorbidity score *i* with a patient having comorbidity score *j*.

For mixing matrices by antibiotic agents, we followed a ranking scheme proposed by RW Moehring et. al.^21^ According to this scheme, antibiotic agents are categorized on a 4-point scale based on their spectrum of activity against bacterial pathogens and priority for antibiotic stewardship program which is as follows: Narrow-spectrum (Rank 1), Broad-spectrum (Rank 2), Extended-spectrum (Rank 3) and Protected (Rank 4). The resulting mixing matrices become 4 × 4 tables where cell [i, j] represents the number of contacts occurring between patient pairs exposed to agent ranks i and j respectively.

Note that the mixing matrices differed for different units within a hospital or for different hospitals. To help with comparisons between different hospital sizes, we also computed a normalized representation for the mixing matrices.

### Software Implementation and Code Availability

Data preparation and network extraction was performed using Python 3.6.9, including the Pandas library for data preparation, and the Bokeh^22^ library for visualization and the creation of interactive plots. These interactive plots allow for comparing and contrasting the mixing matrices across different units, and across different hospitals, and are available at http://go.wsu.edu/hospitalmatrix. Further visualization was conducted in R 3.6.3^23^ using the networkD3 library. The extracted patient contact networks, as well as the source code used for the analysis, are available on at https://github.com/epimodels/mixing_pattern.

## Results

### Patient Contact Networks

In Figure 1, we added a snapshot of the patient contact networks that was generated using the records of one month (January 2017) for each of the hospitals included in this study. The nodes represent patients, and edges represent a pairwise contact between the corresponding two patients in the time interval considered. As expected, most networks were dense, particularly for hospitals with large unit capacities (e.g., Hospital-24, Hospital-14). However, networks were relatively sparse (e.g., Hospital-7, Hospital-16) in smaller hospitals. The largest network in this collection (for Hospital-24) has 9,778 nodes and average degree (the number of edges connecting to that node - in this case the number of co-located patients) of 329; while the smallest network (for Hospital-16) has 572 nodes and an average degree of 35.

**Figure 1:**
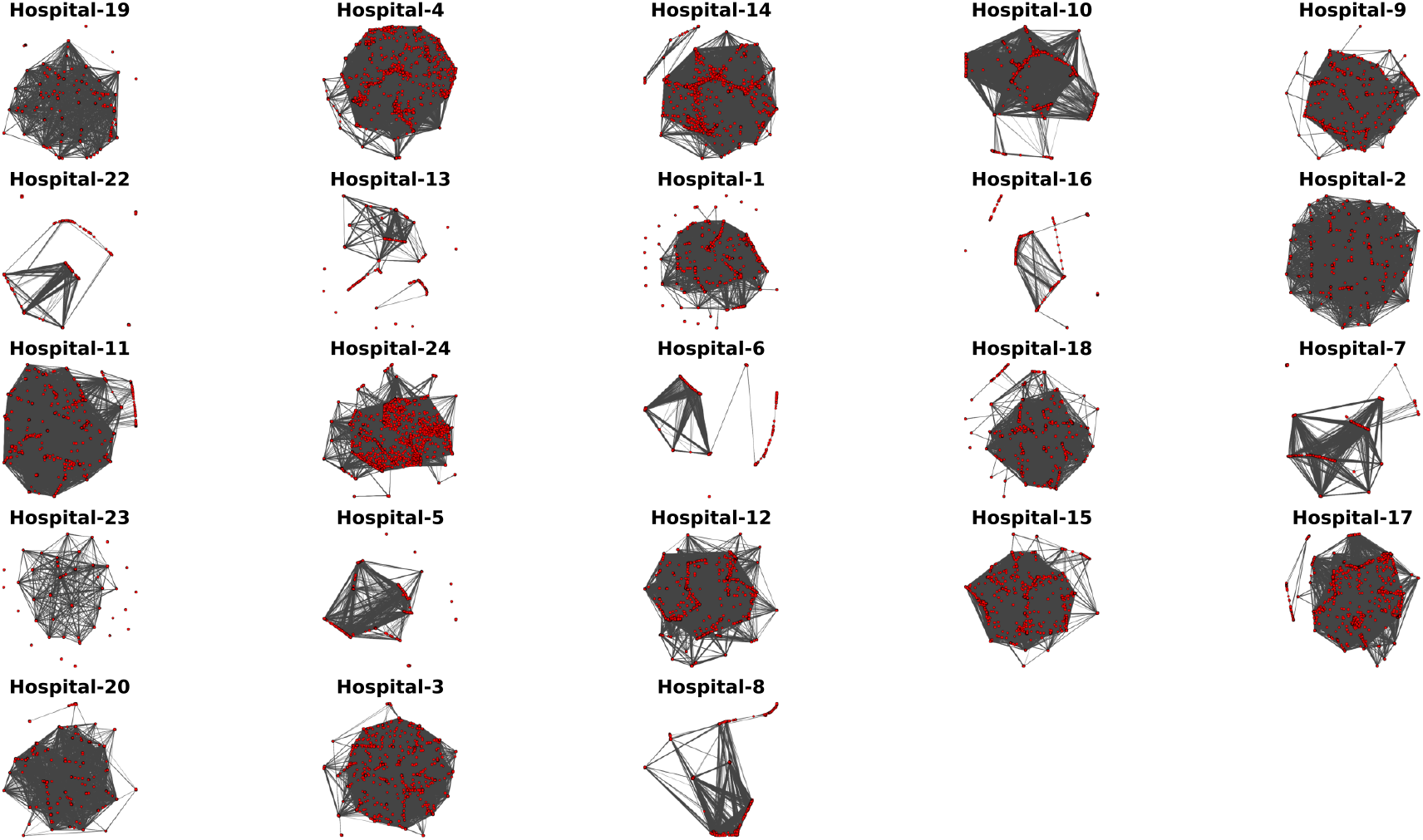
A snapshot of patient contact networks constructed for the DASON data for the month January 2017. Each node is a patient, and each edge represents a contact between the two corresponding patients in that hospital during that month. One hospital is omitted due to a very sparse connectivity over the chosen month.

### Mixing Matrix by Age

There is considerable inter-hospital variability in the age-mixing patterns of patients as shown in Figure 2, owing to the type of hospital, catchment population, etc. There are primarily three major patterns visible: a) hospitals showing uniform mixing across all adult ages (e.g., Hospital-23); b) hospitals serving primarily younger age groups (e.g., Hospital-11); and c) hospitals, especially smaller ones, dominated by mixing between elderly patients (e.g., Hospital-5). These mixing patterns of hospitals were consistent with their respective age distributions (eFigure 1).

**Figure 2:**
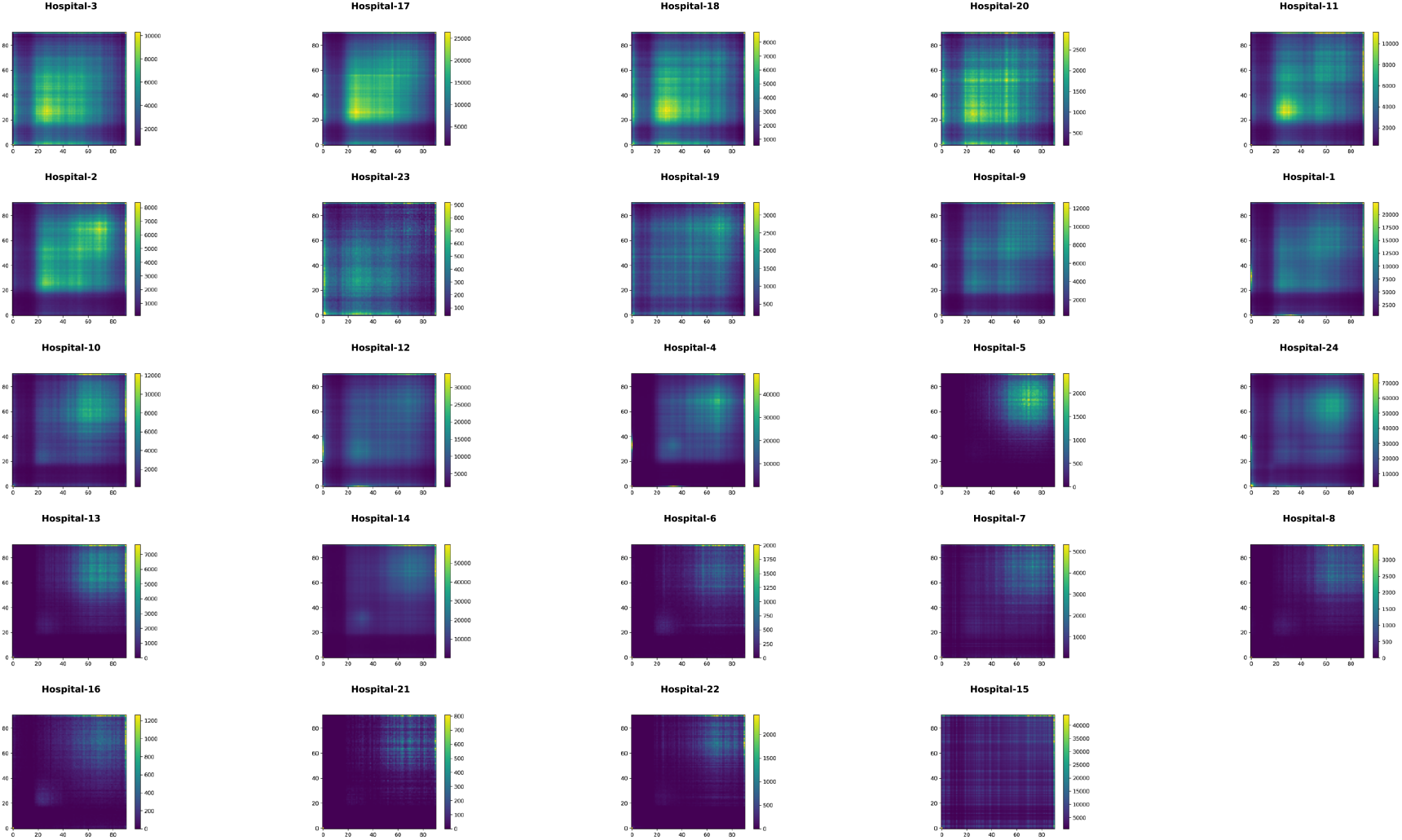
Mixing matrices by age measured by the number of pairwise contacts (shown by the color bar) for each of 24 community hospitals in the DASON network. Brighter areas of color represent denser numbers of patient-to-patient connections based on occupation in the same unit. Each hospital is represented on its own scale.

However, within a hospital, the patterns varied from unit to unit as different units cater to different types of patients. Figure 3A to 3F show a selected subset of six mixing matrices drawn from different hospital units. A darker color shade denotes lower mixing and a brighter color shade denotes heavier mixing.

**Figure 3:**
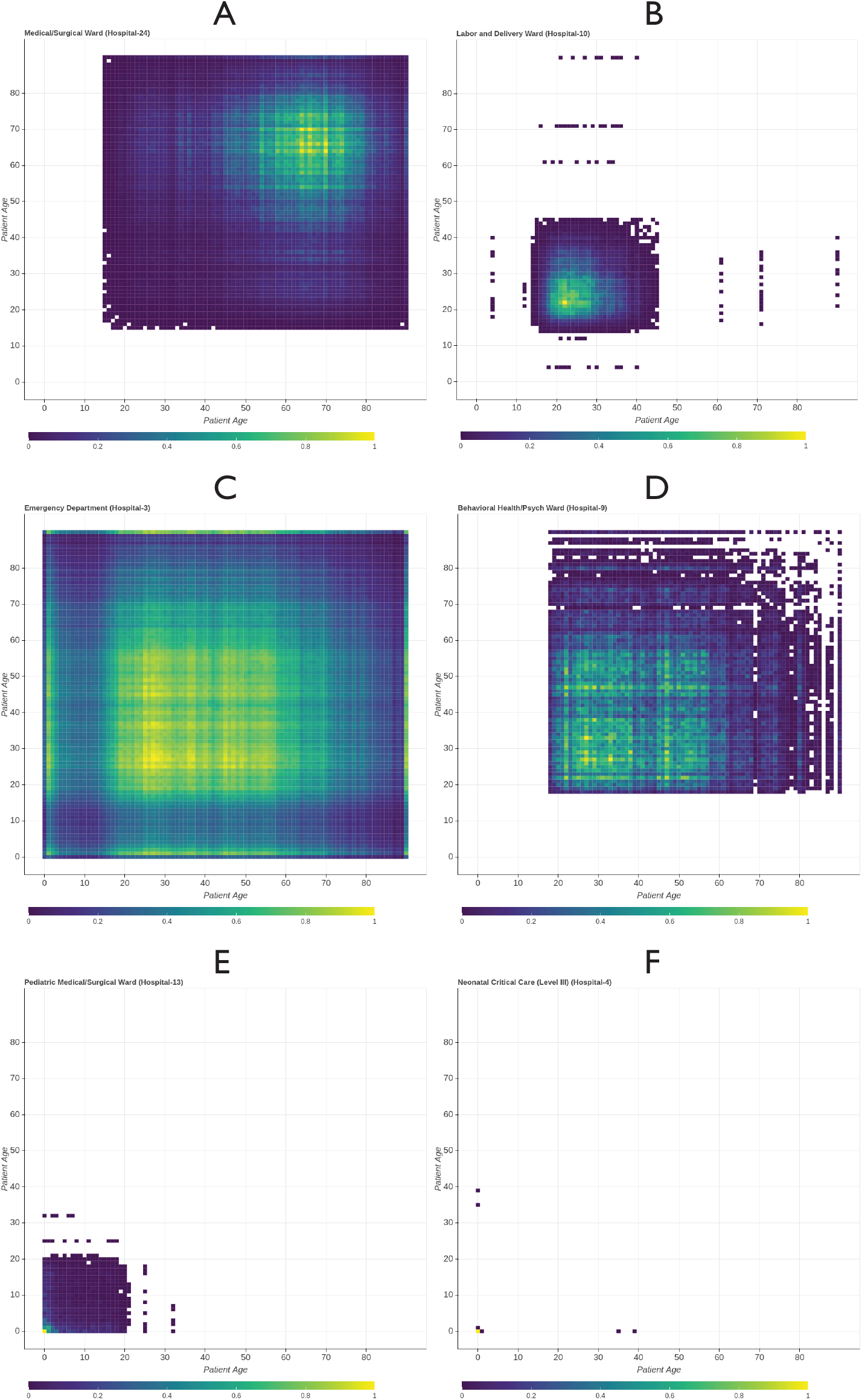
Mixing matrices by age measured by the number of pairwise contacts (shown by the color bar) for selected units in DASON network. Panel A shows a typical adult inpatient unit. Panel B shows a representative pattern of a Labor and Delivery Ward. Panel C is drawn from an Emergency Department. Panel D is from a Behavioral Ward. Panels E and F show the representative patterns in Pediatric and Neonatal units, respectively. Small numbers of incongruous patients (i.e. Labor and Delivery patients 60 years of age or higher) represent small amounts of misclassification, either in patient demographics or the NHSN unit type as a proxy for what a ward is being used for, but are retained for completeness.

Most of the adult inpatient units (e.g. Medical/Surgical - Figure 3A) follows what one might expect - a larger number of patients aged 40 and up, with a high density of patients around 65 years of age, with relatively few young patients. Labor and Delivery wards similarly showed robust patterns between hospitals, concentrated among patients 20 to 35, tapering off toward the limits of maternal age (Figure 3B). The mixing patterns for some other units, such as Operating Room Suites and 24-Hour Observation, showed wide variation between hospitals, representing differing catchment populations and patient mixes. Outpatient units, such as Emergency Departments (Figure 3C) and Behavioral Health (Figure 3D) units had mixing between broager age ranges, with a larger proportion of the mixing between patients in the [20,50] age range. In addition, the ED also showed mixing between adult and child age groups. Pediatric units (Figure 3E) showed broad mixing of young patients under 20, though concentrated in infants. Neonatal units (Figure 3F) are dominated by mixing between infants and young adults of childbearing age.

### Mixing Matrix by Elixhauser Score

Thet Elixhauser Score in DASON data is ranged from 0 to 16, with a median and IQR of 0 due to most patients having lacked known comorbidities. Figure 4 shows the mixing matrices by Elixhauser Score for a selected subset of six different units taken from different hospitals. Patients in adult medical wards generally had mixing between patients with low to moderate Elixhauser scores, with the highest density being patients with a score of four mixing with other patients with a score of four; scores above ten are exceedingly rare (Figure 4A). Adult critical care units (Figure 4B) shifted this distribution up and to the right. Both non-critical (Figure 4C) and critical (Figure 4D) pediatric wards had distributions much more skewed toward patients with zero comorbid conditions, albeit with a broader range of possible values in critical care units. Neonatal units were primarily concentrated in the [0,0] cell. Labor and Delivery Wards (Figure 4E) and Emergency Departments (Figure 4F) had characteristically low rates of comorbid conditions, but the latter demonstrated a wider potential range.

**Figure 4:**
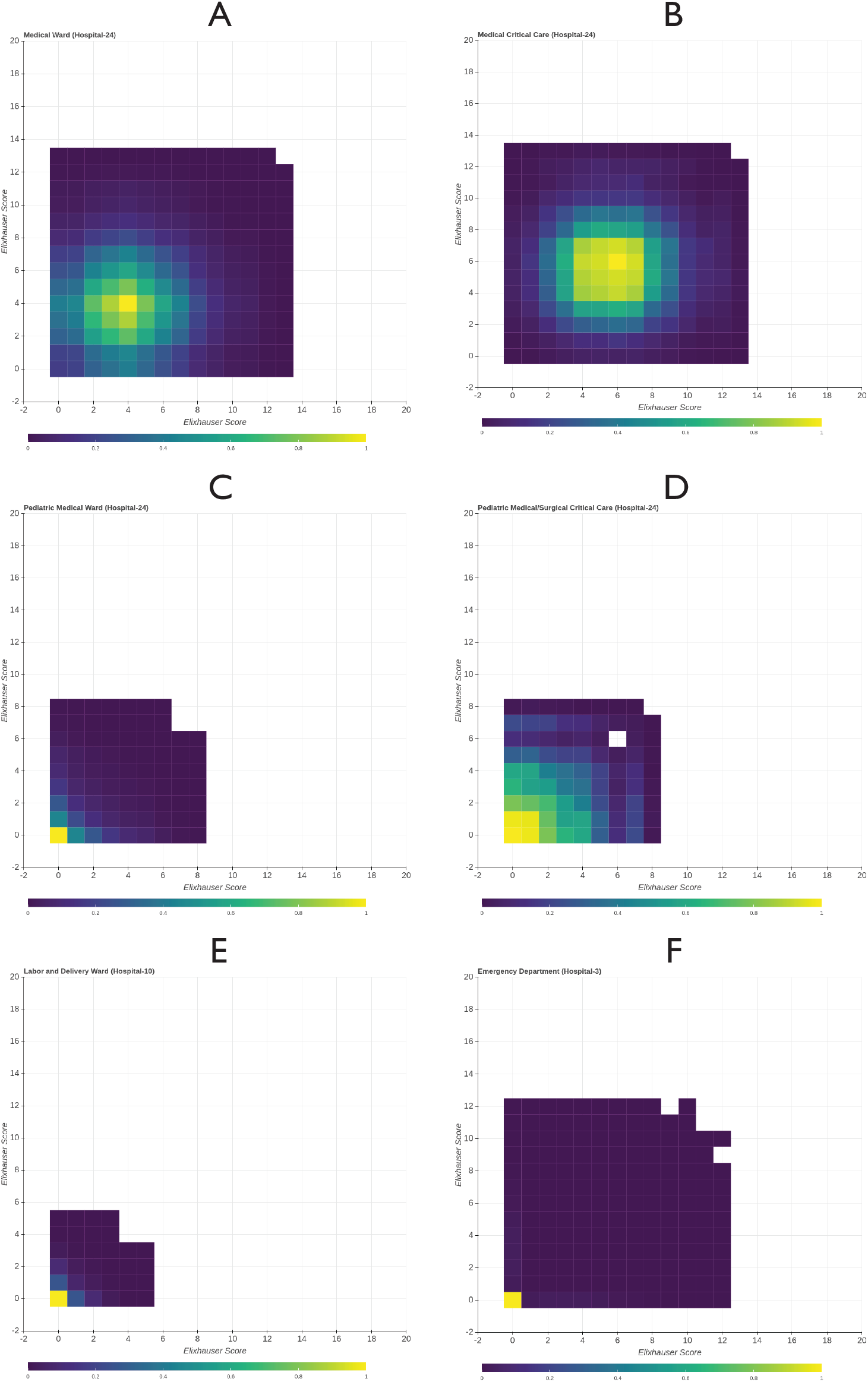
Mixing matrices by Elixhauser Score measured by the number of pairwise contacts (shown by the color bar) for selected units in DASON network. Panels A to D depicts different units within the same hospital, showing an adult medical ward (A), adult ICU (B), pediatric medical ward (C) and pediatric ICU (D). Panel E and F show the patterns for a typical Labor and Delivery Ward, and an Emergency Department respectively.

### Mixing Matrix by Antibiotic Category

From the medication information of DASON data, we found that there were many (and often a large majority) of patients who were not on any form of antibiotic. The ratio of contacts where neither patient in a connected pair were on antibiotics compared to pairs where one patient or both patients were on antibiotics varied widely by unit, shown in eFigure 2 for four selected units.

We observed six distinct patterns on the antibiotic mixing matrices based on the four-point ranking scheme as shown in Figure 5. Across all hospitals, Gynecology, Labor and Delivery and Postpartum units predominantly involved patients on narrow-spectrum agents (Figure 5A). This pattern also occurred in some, but not all, Operating Room Suites and Orthopedic Wards, driven by prophylactic and post-operative cefazolin respectively. Broad-spectrum heavy patterns appeared primarily in Pediatric Medical Surgical Wards (Figure 5B). Wards heavily using extended-spectrum agents (Figure 5C) were most often Adult Critical Care units of all types. This pattern was also observed in some hospitals in Post Critical Care units, Endoscopy Suites, and 24-hour Observation areas. Figure 5D shows a distinctive Narrow-Extended spectrum heavy mixing pattern that appeared predominantly in pediatrics-focused units, including Well Baby Nurseries, Step Down Neonatal Nurseries, and Neonatal Critical Care units and also in a subset of Cardiac Catheterization units, Surgical Cardiothoracic Critical Care units, and Neurological Critical Care units. Broad-Extended spectrum dominated pattern (Figure 5E) was most often seen in Emergency Departments, as well as occasionally in some Telemetry Wards and 24-hour Observation Areas. A final pattern, involving the relatively frequent use of Narrow, Broad and Extended spectrum antibiotics (Figure 5F) was only observed in the Pediatric units of the large academic medical center, discordant with the Pediatric units of the community hospitals.

**Figure 5:**
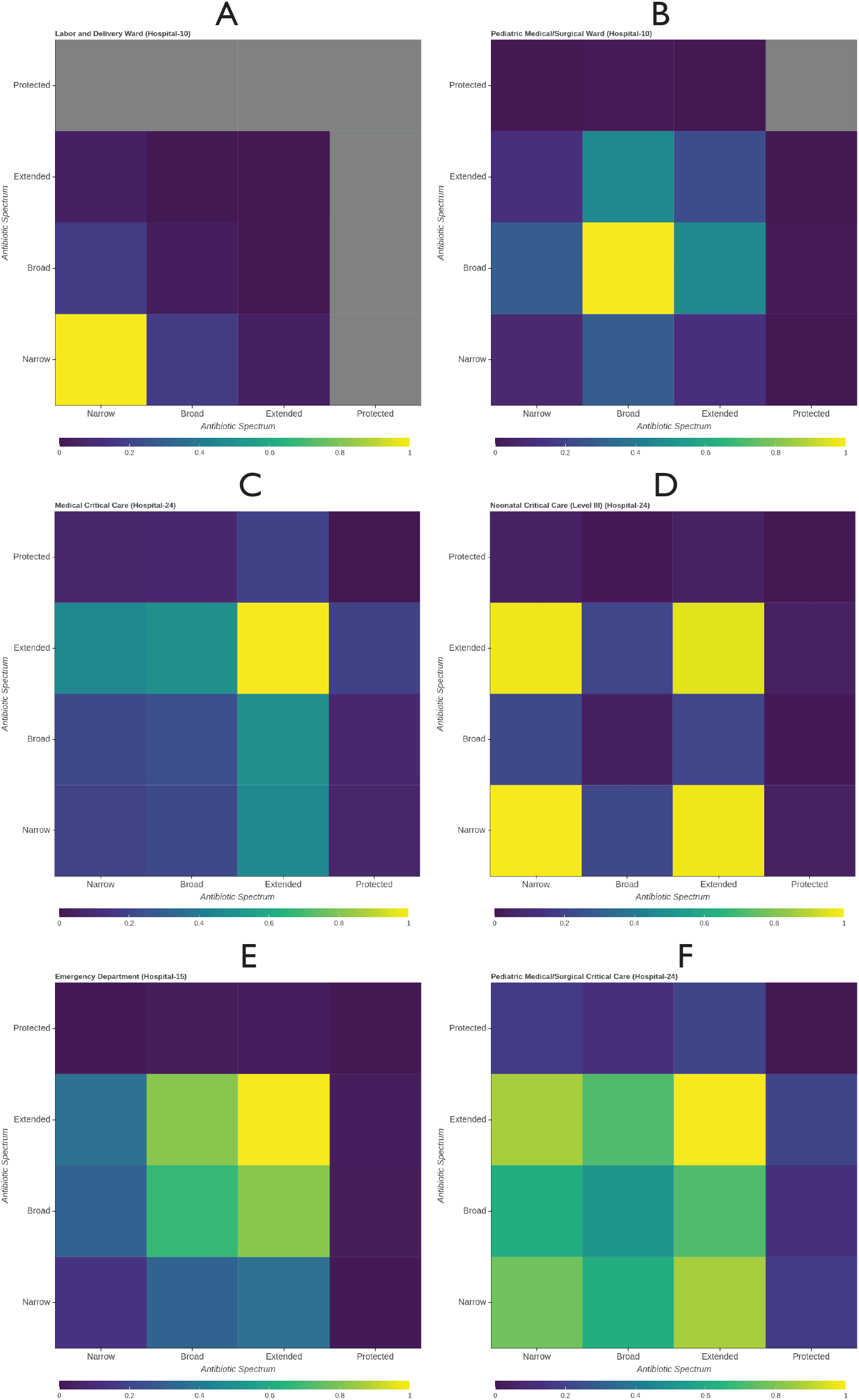
Mixing matrices by antibiotic rank measured by the number of pairwise contacts (shown by the color bar) for selected units in DASON network. Panel A represents a unit where the patients who came in contact were mostly exposed to one type of antibiotic namely Narrow spectrum antibiotics. Panels B and C show Broad and Extended spectrum heavy contact patterns. Panels D and E show highlighted inter-spectrum mixing patterns, e.g. Narrow-Extended and Broad-Extended. Panel F highlights a wide region encompassing Narrow-Broad-Extended spectrum antibiotics.

## Discussion

### Mixing Matrix by Age

Mixing matrices by age mostly conform to the expected patterns one would expect heuristically with a few exceptions. Especially Emergency Departments and Behavioral Units showed areas with broader age-related mixing patterns. These units require special consideration when considering pathogens with markedly different age-related risks, transmission potentials, or vaccination status.

### Mixing Matrix by Elixhauser Score

In comparison to general units, the dense area in the critical care units shifted up and to the right, indicating -- not unexpectedly -- an increase in patients with more prevalent comorbid conditions, though the peak for this distribution was more diffuse. Patients in the pediatric and neonatal units are not likely to have developed comorbid conditions which explains the dense region on the [0,0] cell although pediatric critical care has a broader range of possible values reflecting their more complex patient mixture. The Emergency Department, although concentrated on the zero comorbid zone, demonstrated mixing between a wide potential range reflecting its central role as a possible place for patients with vastly different underlying characteristics to encounter one another.

### Mixing Matrix by Antibiotic Category

Mixing matrices that showed dense mixing on one rank is an indicator of substantial use of a specific kind of antibiotic in a unit. On the other hand, the bimodal inter-spectrum mixing can arise from one of two possible mechanisms -- two distinct groups of patients, one on one type of antibiotic, the other on another type of antibiotic, who happened to be co-located in the same unit. The second is that the same patients are prescribed drugs of two different ranks. To examine these two possibilities, we considered one ward with both a large number of patients as well as the distinctive Narrow-Extended spectrum pattern, a neonatal intensive care unit in a large academic medical center. The distribution of antibiotic exposures are shown in eTable 1. It is apparent that the vast majority of patients are exposed to both classes of antibiotics during their hospitalization, though the result was not statistically significant (Fisher’s Exact Test *p* = 0.056). This pattern seems likely to arise from a commonly used combination of ampicillin and gentamicin for empiric coverage of neonatal sepsis^24^. This, in turn, suggests that a patient’s individual antibiotic exposure profile likely represents their primary exposure at any given time.

In contrast, the analysis of a unit (Emergency Department in Hospital-15) having a Broad-Extended pattern, reveals a different picture as can be seen from the distribution in eTable 2. This unit served a large elderly population with several Skilled Nursing Facilities, and among them 77.6% of patients were prescribed either broad- or extended-spectrum agents (33.2% and 44.4% respectively) (Chi-squared *p* > 0.001). These findings once again highlight the importance of Emergency Departments as areas with far broader mixing patterns than the rest of the hospital environment, as well as the likelihood of the units discussed above being central points of empiric therapy within many hospitals.

The inter spectrum mixing of Narrow, Broad and Extended spectrum antibiotics was only observed in the Pediatric units of the large academic medical center. The academic medical center is notable for having both Cystic Fibrosis patients as well as a pediatric transplant program, which resulted in a markedly different patient profile to community hospital Pediatric units, and likely drove these different patterns.

## Conclusions

This study presents several aspects of how hospitalized patients come into contact with each other. Understanding these contact patterns can provide vital information on infection transmission risk - for example, where patients with high potential susceptibility to acquiring multidrug resistant organisms might be in contact, directly or indirectly, with those at serious risk for adverse outcomes from infection. While some of these patterns may be inferred heuristically, it is nevertheless beneficial to quantify these patterns. This enables their use in modeling studies of hospital-acquired pathogens. Further, it could provide a means to quantitatively track shifts in antibiotic use or patient case mix patterns. For example, a unit shifted away from its NHSN unit-type designation to deal with surges of COVID-19 patients, and the population characteristics changed. As another example, the matrices might change as stewardship programs intervened on how antibiotics were used. Finally, quantifying the variability *between* hospitals can help assess the generalizability of effect estimates and the ensuring intervention and policy recommendations between the academic medical centers where these estimates are often obtained and rural and community hospitals, for any situation where patient-to-patient interaction is potentially at play.

There are important limitations to this study, arising from the use of an existing data source to reach multiple hospitals and a large number of patients. This study implicitly assumes that patients visiting a unit the same day had contact - primarily via indirect contact mediated by either healthcare workers or the environment. While age, underlying comorbidities and antibiotic exposure are certainly important risk factors for healthcare associated infections, they are far from an exhaustive list, and there are a number of risk factors that are beyond the reach of a single study of this type, or impractical to collect on an ongoing and continual basis in a broad network of hospitals of varying resource levels. Besides, patients that occupy the same unit for multiple days have repeated contacts, which we assume linearly add to the amount of mixing. Finally, we assume that an NHSN unit designation is an adequate proxy for the type of procedures and patients present in a given unit, which may result in a degree of misclassification.

Additionally, a number of contact patterns were either more diffuse (in the case of age and Elixhauser score) or unique (in the case of some antibiotic prescribing patterns in pediatric units) to the academic medical center present in the data set. This highlights the need for data and parameter estimates from both academic medical centers and community hospitals to better inform mathematical modeling studies, estimates of intervention effectiveness, and other studies that rely on the generalizability of estimates from studies that often take place in large academic or tertiary-care hospitals. This study provides a wealth of estimates from hospitals of diverse sizes, catchment areas and patient populations, albeit within a limited geographic area. This should serve to help improve the quality of future studies in this arena.

## Supporting information

eFigure 1

## Data Availability

The extracted patient contact networks, as well as the source code used for the analysis, are available on at https://github.com/epimodels/mixing_pattern.

## Acknowledgments

This research was supported by contract 75D30121P10551 from the US. Centers for Disease Control and Prevention (CDC) and grant DBI 1661348 from the U.S. National Science Foundation (NSF).

