## Supplementary material for "Who Is Hospitalized With Whom? Inpatient Contact Networks and Mixing Patterns": eFigure 1

### SUPPLEMENTARY MATERIALS

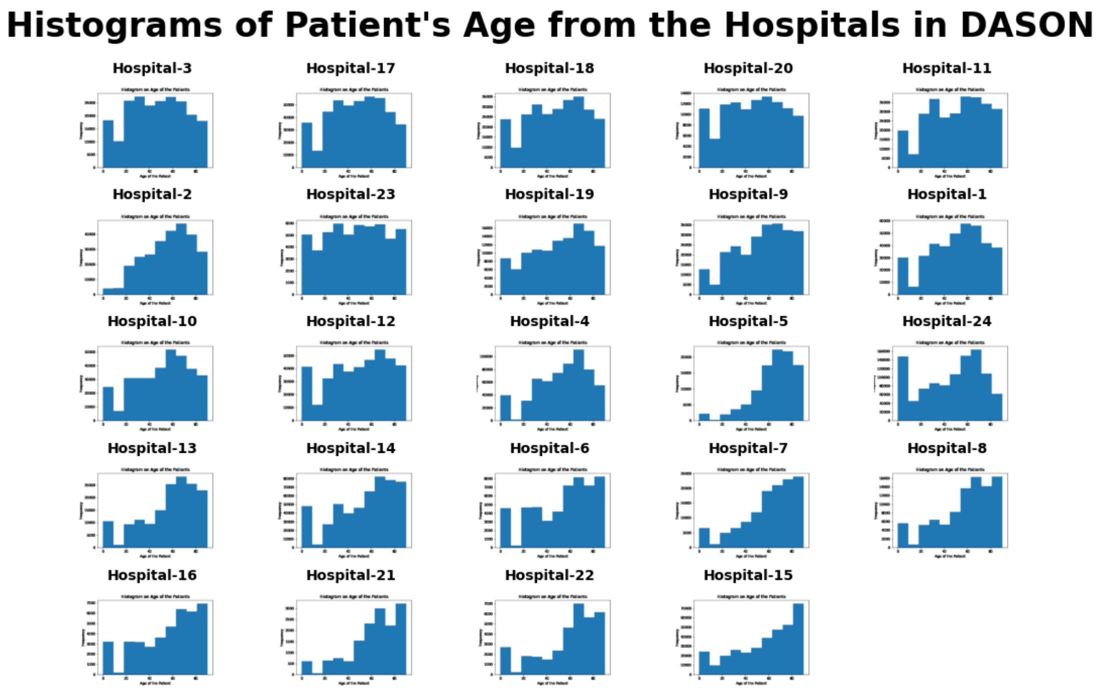


**eFigure 1:** Histograms depicting the distributions of the patient's age who visited the hospitals included in the DASON data set (Time frame: October 2015 to November 2017).

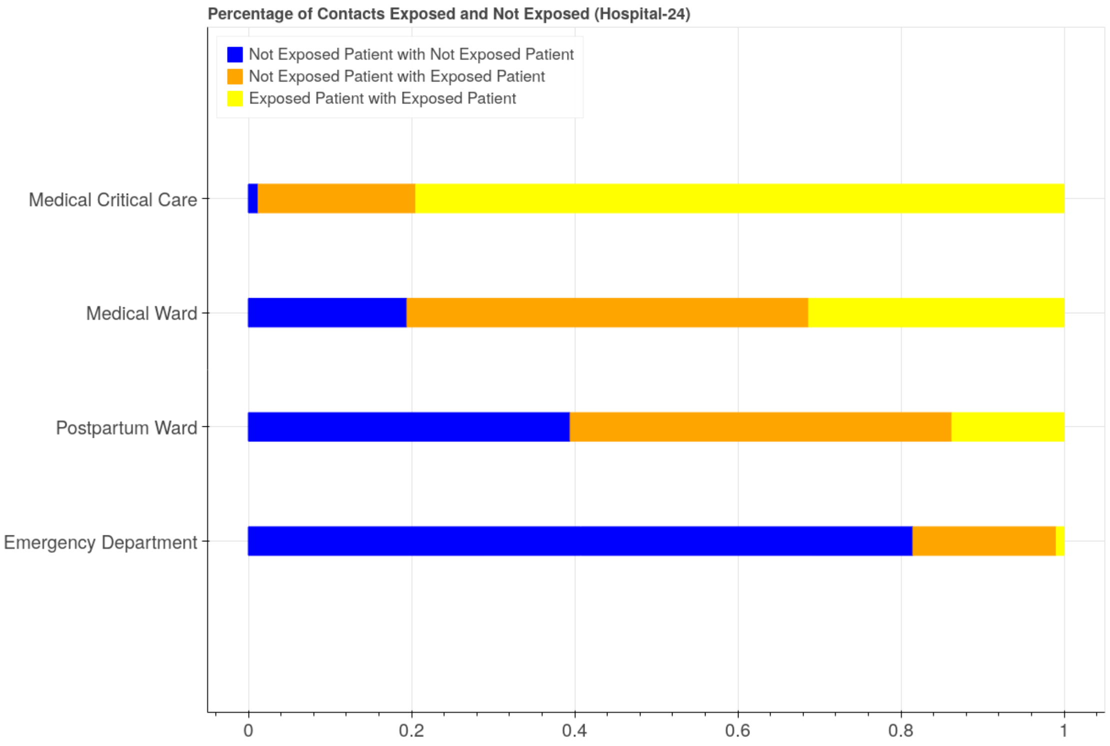


**eFigure 2:** Bar chart depicting the ratio of contacts where the contacts are categorized as (i) No antibiotic for both of the patients who contacted, (ii) One of the patients was on antibiotic and another was not and (iii) Both of the patients were on antibiotic. Here, the top two barcharts show a comparative scenario in a Medical Ward and a Medical Critical Care from the same hospital. While medical care has a higher ratio on one of the patients on antibiotics, the critical care is apparently higher in the ratio for both of the patients on antibiotics. The barchart from Postpartum Ward shows the ratio where almost half of the contacts were between patients who were not on any antibiotic and for the remaining half is distributed amongst the other two categories. And the last is drawn from the Emergency Department which shows most of the contacts occurred in this unit were between patients who were not exposed to any antibiotic until that point.

**eTable 1. Distribution of patients (n=1285) in a Neonatal Critical Care (Hospitalid 2000) shown based on their exposure to Narrow and/or Extended spectrum antibiotics.**

| | Narrow-Spectrum Exposed | Narrow-spectrum Unexposed |
|---|---|---|
| Extended-Spectrum Exposed | 1,230 | 9 |
| Extended-Spectrum Unexposed | 44 | 2 |

**eTable 2. Distribution of patients (n=14633) in an Emergency Department (Hospitalid 1045) shown based on their exposure to Broad and/or Extended spectrum antibiotics.**

| | Took Broad-spectrum | Did not take Broad-spectrum |
|---|---|---|
| Took Extended-spectrum | 2,445 | 6,499 |
| Did not take Extended-spectrum | 4,856 | 833 |
